# Examine the impact of weather and ambient air pollutant parameters on daily case of COVID-19 in India

**DOI:** 10.1101/2020.06.08.20125401

**Authors:** Kousik Das, Nilanjana Das Chatterjee

## Abstract

The present study presents a view on exploring the relationship pattern between COVID 19 daily cases with weather parameters and air pollutants in mainland India. We consider mean temperature, relative humidity, solar radiation, rainfall, wind speed, PM_2.5_, PM_10_, SO_2_, NO_2_ and CO as independent variable and daily COVID 19 cases as dependent variable for 18 states during 18^th^ march to 30^th^ April, 2020.After dividing the dataset for 0 to 10 day, 10 to 25 days and 0 to 44 days, the current study applied Akaike s Information Criteria (AIC) and Generalized Additive Model (GAM) to examine the kind of relationship between independent variables with COVID 19 cases. Initially GAM model result shows variables like temperature and solar radiation has positive relation (*p*<0.05) in 0 to 10 days study with daily cases. In 25 days dataset it significantly shows that temperature has positive relation above 23 degree centigrade, SO_2_ has a negative relationship and relative humidity has negative (between 30% to 45% and > 60%) and a positive relationship (45% to 60%) with COVID 19 cases (*p*=0.05). 44 days dataset has six parameters includes temperature as positive, relative humidity as negative (between 0 to 45%) and then positive (after >45%), NO_2_ as Positive (0 to 35 microgram/m^3^) followed by negative trend (after > 40 microgram/m^3^), SO_2_ and rainfall as negative relation. After sensitive analysis, it is found that weather variables like relative humidity, solar radiation and rainfall are more sensitive than temperature and wind speed. Whereas pollutants like NO_2_, PM_2.5_, PM_10_ and CO are more sensitive variables than SO_2_ in this study. In summary this study finds temperature, relative humidity, solar radiation, wind speed, SO_2_, PM_2.5_, and CO may be important factors associated with COVID 19 pandemic.

**Graphical Abstract:** 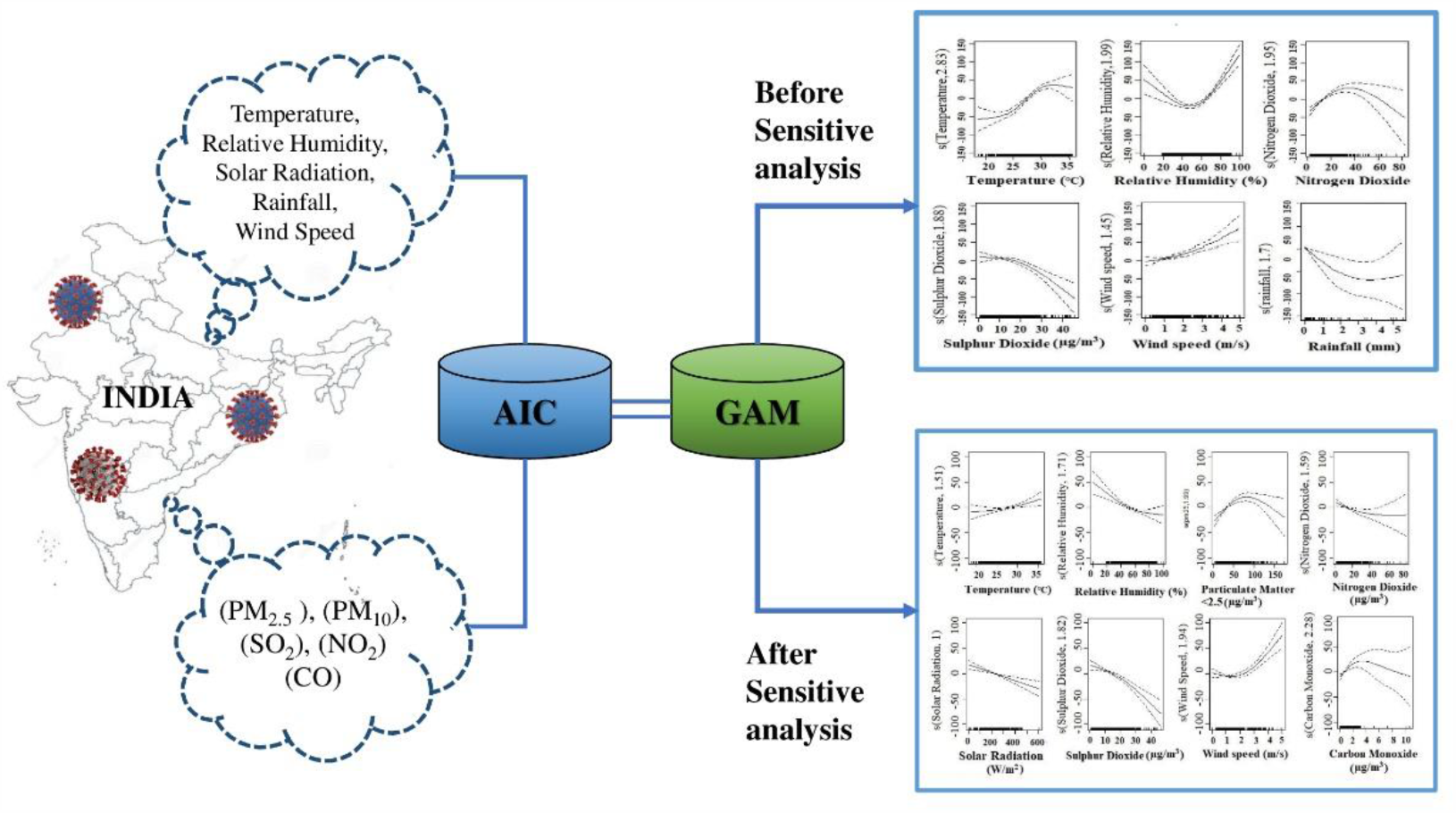

**Highlights:**

➢ There was a significant relationship between daily positive COVID-19 case with weather and pollution factors
➢ We found PM_2.5_ and CO positively associated with transmission of positive cases where as NO_2_ and SO_2_ have a negative relation after sensitive analysis.
➢ We have found temperature and wind speed have positive relation whereas, relative humidity and solar radiation have negative relation after sensitive analysis.
➢ Weather variables like relative humidity and solar radiation and rainfall are more sensitive than temperature and wind speed. Pollutants like NO_2_, PM_2.5_, PM_10_ and CO are more sensitive variables than SO_2_ in this study.

## 1. Introduction

The severe acute respiratory coronavirus 2 (SARS-CoV-2) (Gorbalenya, 2020) is an infectious disease, initially found in Whan, China (Li et al., 2020). Health Organization (WHO) has declared it pandemic worldwide on 11^th^ March, 2020, after spreading to several countries (Cucinotta et al., 2020). COVID-19 infected patients have some typical symptoms including fever, myalgia, dry cough, pneumonia and throat sore (Şahin, 2020. Huang et al., 2020). As of 16^th^ may, 2020, a total 4,425,485 confirmed cases with 302,059 death has been registered so far (https://covid19.who.int/). Where United States of America (USA) is leading with 1.47 million conformed cases followed by Russia, United Kingdom, Spain and Italy. In India total active cases 53035 with 2752 death and 30,152 recovery reported till date of 16^th^ may 2020 (https://www.mohfw.gov.in/). These official data shows that recovery rate 35% and fatality stand at 3.2% in India. Being a densely populated area, India has alluring ground to be affected faster than other countries. But timely management and strict lockdown measures were able to reduce its predicted growth rate as per world level (Gupta et al., 2020).

In addition to human contact, there are many research works seeks to find the relationship of weather factors (Tosepu et al., 2020, Zhu et al., 2020 and Shi et al., 2020), air pollutants (Martelletti et al., 2020, Ogen, 2020 and Conticini et al., 2020), geographical factors (Gupta et al., 2020) and socio-economic factors (Qiu et al., 2019) with daily conform COVID-19 active cases as well as death due to this disease. Weather parameters like Temperature, Relative Humidity, Solar radiation, Rainfall, Wind speed (Chaudhuri et al., 2020 and Zhu et al., 2020) may enhance the transmission of corona positive cases. In addition to this ambient air pollutants like PM_2.5_, PM_10_, SO_2_, NO_2_ and CO (Bashir et al., 2020 and Yongjian et al.,) also could be associated with enhancing the process of COVID-19 cases. However recent study focuses on single or two factor based study like Temperature, relative humidity, solar radiation, SO_2_and Particulate matter etc. Pollutants and weather parameters both have combine effect on cardiovascular and respiratory disease (Delamater et al., 2012 and Vanos et al., 2014). Since sever acute respiratory coronavirus 2 (SARS-CoV-2) causing chronic damage and injury to the cardiovascular system (Zheng et al., 2020) it may have positive association with high concentration of Particulate Matter (PM) (Yongjian et al., 2020). Again meteorological condition significantly influenced the concentration of PM (Zhao et al., 2014). Therefore we assume that combine effect of weather parameters and pollutants will be a holistic way to find associative factors. The major objectives is to explore the relationship of weather parameters and air pollutants with COVID-19 cases. Unlike other study, we consider a state level rather than one city. Considering 14 days of moving average for temperature and relative humidity, each parameters were divided into three timeframe i.e. 1 to 10 days, 1 to 25 days and 1 to 44 days. This way of study will provide a holistic insight to study relationship of weather air pollutants with daily corona positive cases.

## 2. Material and Method

### 2.1 Study Area

India is country with 3,287,240 sq. km of its geographical area cherished a unique type of climate throughout of the year. To study the relationship between spread of COVID 19 cases with weather parameter and pollutants over mainland of India, this study comprise a state level experiment includes 18 Indian states which covered most of the portion of India. The states having a considerable number of COVID cases recorded till 30^th^ April of 2020, have been taken for study these are Andhra Pradesh, Assam, Bihar, Delhi, Gujrat, Haryana, Jharkhand, Karnataka, Kerala, Madhya Pradesh, Maharashtra, Punjab, Rajasthan, Uttarakhand, Tamil Nadu, Telangana, Uttar Pradesh, and West Bengal. This kind of study appeal for micro level experiment and because of unavailability of meteorological station data lead to make it study on state level.

### 2.2 Data Collection

Daily lab tested report of COVID 19 of 18 states has been collected from the Ministry of Health and Family Welfare (MoHFW) since seven days prior to first lock down day on 18^th^ March to 30 April, 2020. Meteorological and air pollutants data like daily mean temperature, relative humidity, solar radiation, rainfall, wind speed and Pollution data like mean daily concentration of Particulate Matter less than 2.5 micrometre (PM_2.5_), Particulate Matter less than 10 micrometre (PM_10_), Sulphur Dioxide (SO_2_), Nitrogen Dioxide (NO_2_) and Carbon Monoxide (CO) have been collected from Central Pollution Control Board (CPCB) website. A common issue is CPCB weather stations are not equally distributed all over India therefore mean values of all stations was calculated within a state is considered for final value of that state. Some of the states have fewer inactive stations that would again simulated by nearest station in neighbour state in respect to same physio-climatic condition.

### 2.3 Statistical Analysis

A Generalized Additive Model (GAM) is a combine form of Generalized linear Model (GLM) and Additive Model (AM). GLM has linear predictor acts with sum of covariant smoothing function (Ravindra et al., 2019) on the other hand GAM allows the special additive and non-linear function of the variables. In this study, the GAM along with Gaussian distribution family, is used to examine the non-linear relationship on COVID-19 positive case with weather and pollution parameter. It is considered to be useful to study the nonlinear pattern of relationship lied in between health effect and weather factor (Talmoudi et al., 2017, Cheng e.t al., 2014 and Gasparrini at al., 2010). A moving average approach has been inculcated for better understating of cumulative lag effect of temperature and relative humidity on positive COVID-19 cases (Zhu et al., 2020, Yongjian et al., 2020 and Jan et al., 2007).

It is better choice to use 1-14 days of moving average in temperature and relative humidity because of the incubation period of COVID-19 range for 0 to 14 days (Zhu et al., 2020, Duan et al., 2019 and Lu et al., 2015). Additionally smoothing spine functions of time (Ma et al., 2020, Zeng et al., 2016 and Cheng et al., 2012) has been incorporated to coincide monotonic and non-monotonic pattern among COVID-19 cases with weather and pollution parameter offers a more flexible model. AIC (Akaike’s Information Criteria) was use as a measure for selecting those variables having some kind of association exist in between COVID-19 case and the parameters are used in this study to encounter model overfitting (Yen et al, 1998).

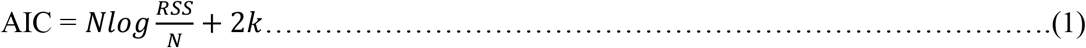

Where N is the number of observation, RSS is the residual sum of square, k is the number of parameters to fit plus 1 (*k* = β+1).

AIC selects the optimum number of variables on the basis of changes in R-squared in a multivariate regression. Smaller AIC value is needed to select the best model (Posada et al., 2001). We apply a step wise regression by considering both forward and backward direction method in R studio environment to identify variables with significant correlation with COVID cases. After selecting variables we further feed them into GAM model, defined as:

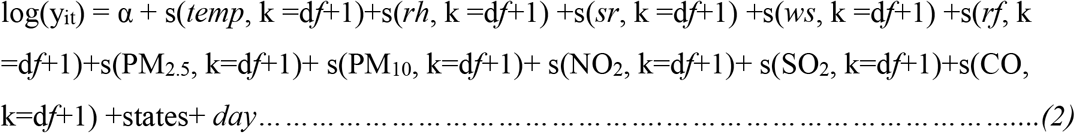

Where log(y_it_) is the log-transformation daily conform cases of COVID-19 on the day t; α indicates intercept; Regression coefficient is designated as β; (*s*) demarks smoothing function based on penalized smoothing spline. Based on previous literature and AIC result we used 2 degree of freedom (*df*) for time and 2-3 for other variables (Zhu et al., 2020 & Ma et al., 2020) for avoiding overfitting (Wang et al., 2018).

We also executed three way of sensitive analysis by dividing our total data set into 3 part (0-10 days, 0-25 days and 0-44days of lockdown). Another way of sensitive analysis is performed by changing degree of freedom (2 – 9 *df* for time and other variable) in model execution time (Ma et al., 2020). And most importantly repeat the model by excluding Maharashtra state as it has large number of case concentrate in last couple of days (Zhu et al., 2020).

Since the model considers a relationship of weather parameters and pollutants with COVID cases, effect plots having visual interpretation of log transform daily case with other variables w ere produced. Significance level of 95% was adopted for all statistical analysis. Whole analysis were performed in R software (version 4.0.0) with the help of “mgcv” package (version 1.8-31). For mapping purpose we have used ArcGis software (version 10.4.1). Spearman’s correla tion has calculated to see the significance correlation among the variables of meteorological, pollutant and COVID-19 cases.

## 3. Result

### 3.1 Description of daily positive case, meteorological parameters and pollutants

Table 1 describes the general information about daily positive case, Meteorological parameters and pollutants, during the study time period of 18^th^ March to 30^th^ April. On an average India experienced daily approximately 42 corona positive case with 26 death. Average daily mean temperature, solar radiation, rainfall, relative humidity, wind speed were 27.678 °C, 227.7W/m^2^, 0.096 mm, 55.95 % and 1.342 m/s respectively. Whereas average mean concentration of Particulate Matter less than 2.5 micrometre (PM_2.5_), Particulate Matter less than 10 micron (PM_10_), Sulphur Dioxide (SO_2_), Nitrogen Dioxide (NO_2_) and Carbon Monoxide (CO) were 38.22 µg/m^3^, 73.977 µg/m^3^, 13.075 µg/m^3^, 15.43 µg/m^3^ and 0.819 µg/m^3^(Fig. 3).

**Table 1.**
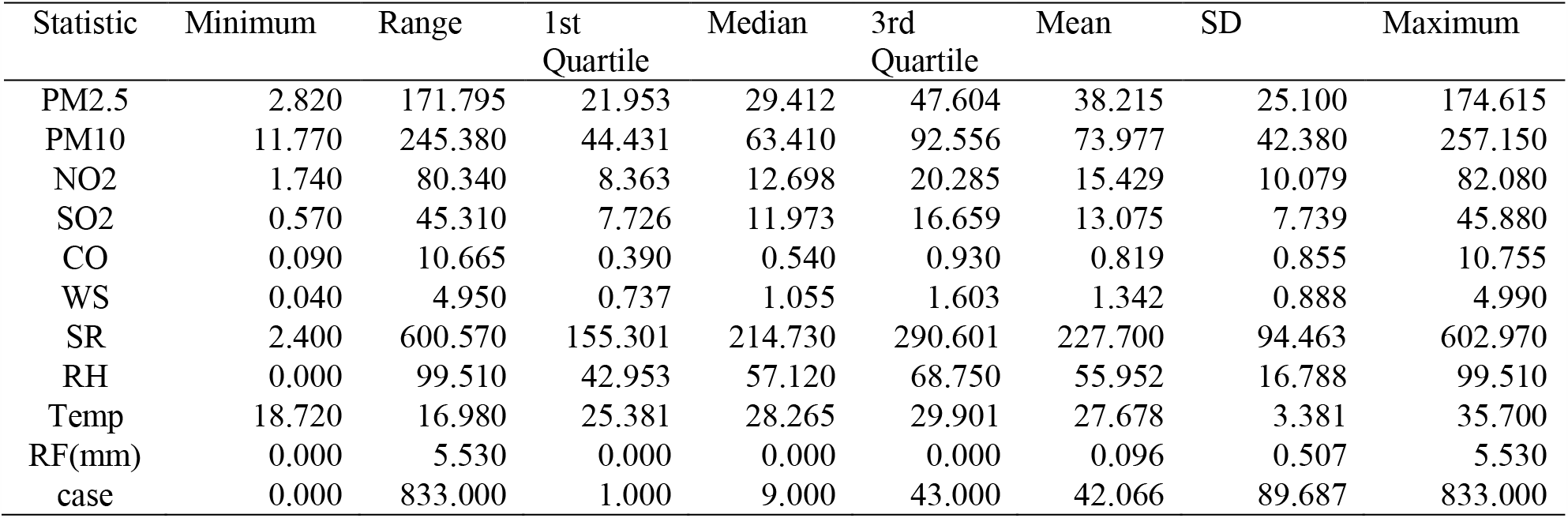
Descriptive statistics of daily conformed COVID cases, meteorological and air pollutants

| Statistic | Minimum | Range | 1st Quartile | Median | 3rd Quartile | Mean | SD | Maximum |
| --- | --- | --- | --- | --- | --- | --- | --- | --- |
| PM2.5 | 2.820 | 171.795 | 21.953 | 29.412 | 47.604 | 38.215 | 25.100 | 174.615 |
| PM10 | 11.770 | 245.380 | 44.431 | 63.410 | 92.556 | 73.977 | 42.380 | 257.150 |
| NO2 | 1.740 | 80.340 | 8.363 | 12.698 | 20.285 | 15.429 | 10.079 | 82.080 |
| SO2 | 0.570 | 45.310 | 7.726 | 11.973 | 16.659 | 13.075 | 7.739 | 45.880 |
| CO | 0.090 | 10.665 | 0.390 | 0.540 | 0.930 | 0.819 | 0.855 | 10.755 |
| WS | 0.040 | 4.950 | 0.737 | 1.055 | 1.603 | 1.342 | 0.888 | 4.990 |
| SR | 2.400 | 600.570 | 155.301 | 214.730 | 290.601 | 227.700 | 94.463 | 602.970 |
| RH | 0.000 | 99.510 | 42.953 | 57.120 | 68.750 | 55.952 | 16.788 | 99.510 |
| Temp | 18.720 | 16.980 | 25.381 | 28.265 | 29.901 | 27.678 | 3.381 | 35.700 |
| RF(mm) | 0.000 | 5.530 | 0.000 | 0.000 | 0.000 | 0.096 | 0.507 | 5.530 |
| case | 0.000 | 833.000 | 1.000 | 9.000 | 43.000 | 42.066 | 89.687 | 833.000 |

**Fig 1.**
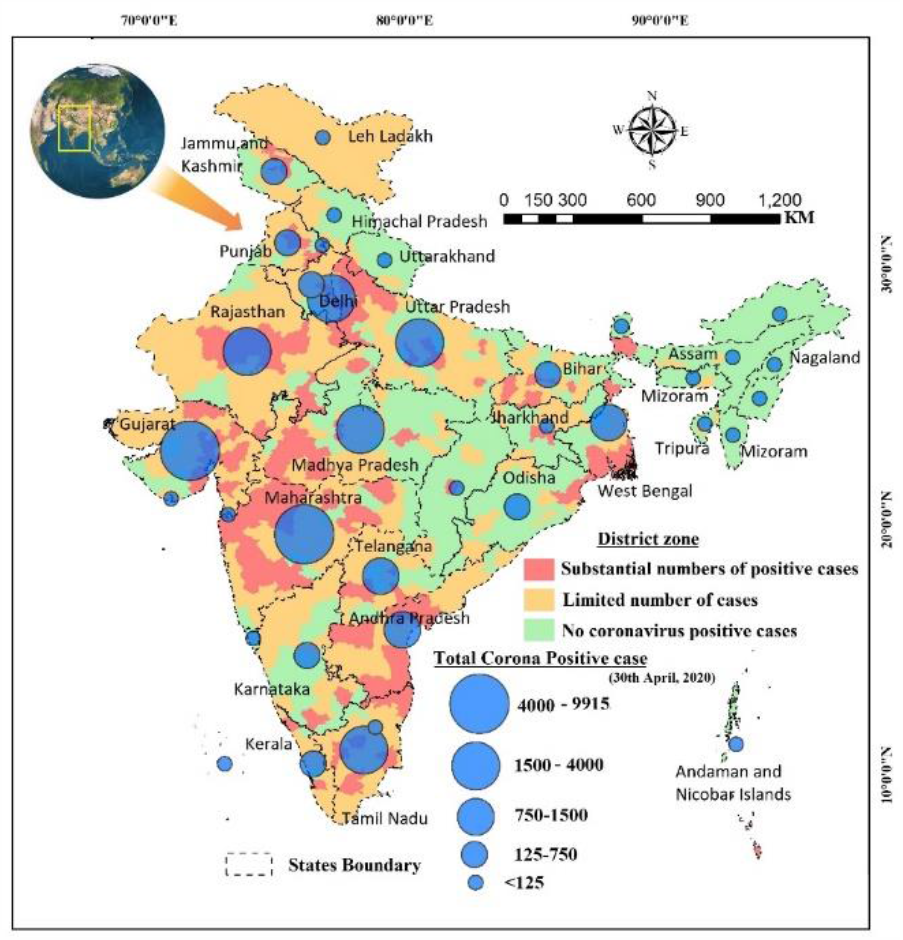
Study area and all states of India and their COVID-19 cases from 18^th^ March to 30^th^ April, 2020.

**Fig 2.**
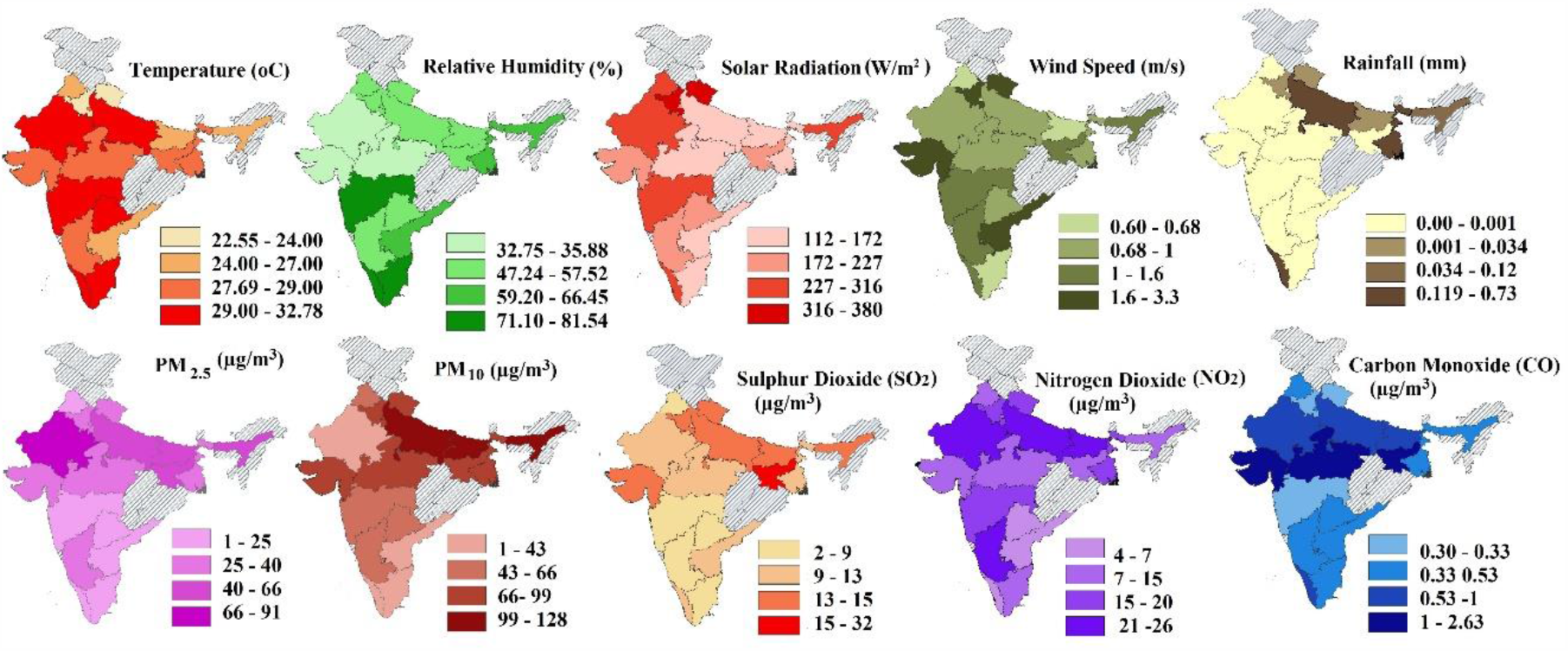
mean value of meteorological parameter and concentration of pollutants from 18^th^ March to 30^th^ April, 2020.

**Fig 3.**
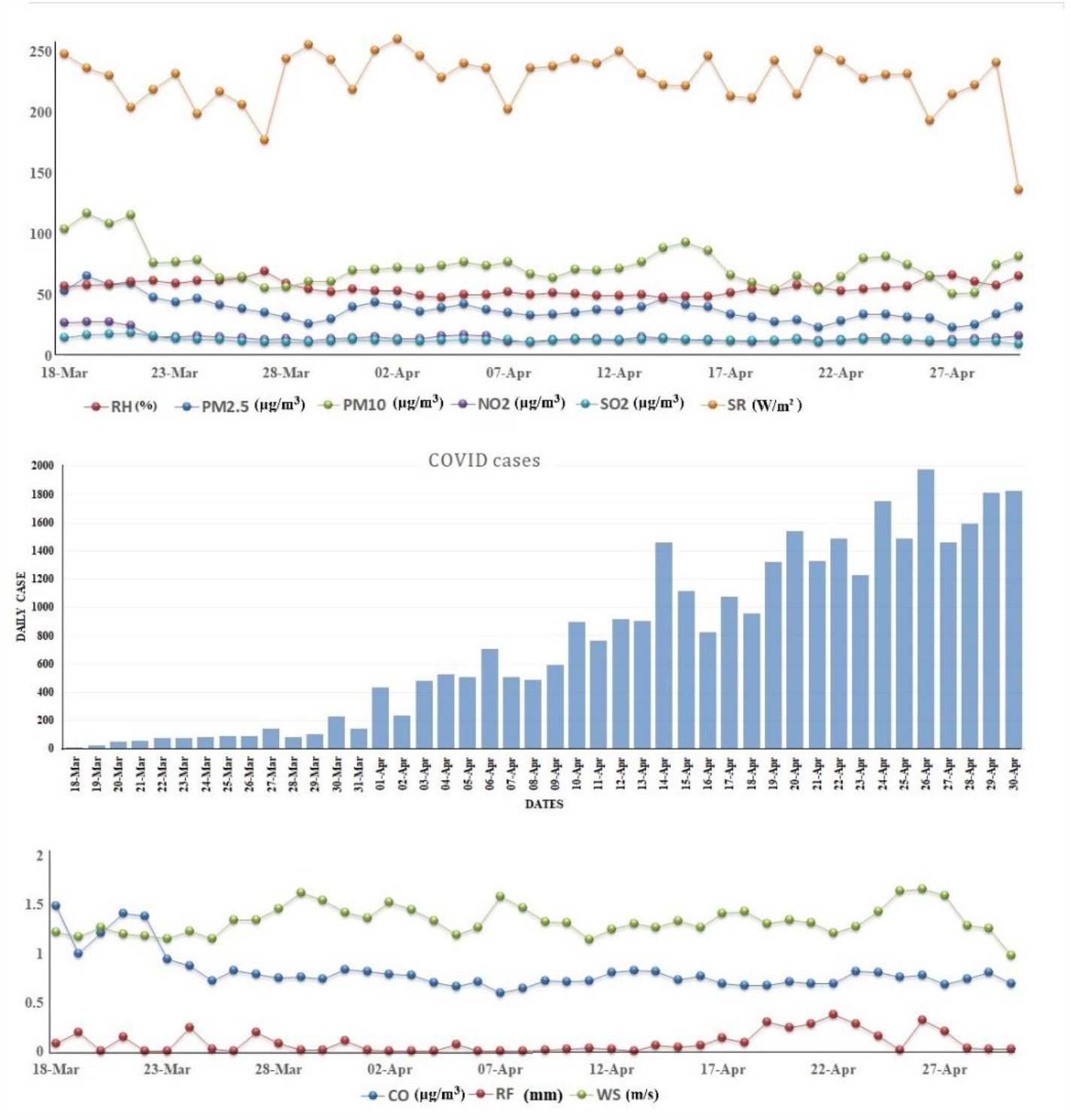
Temporal distribution of daily COVID-19 cases and mean value of weather parameters and pollutants.

**Fig. 4.**
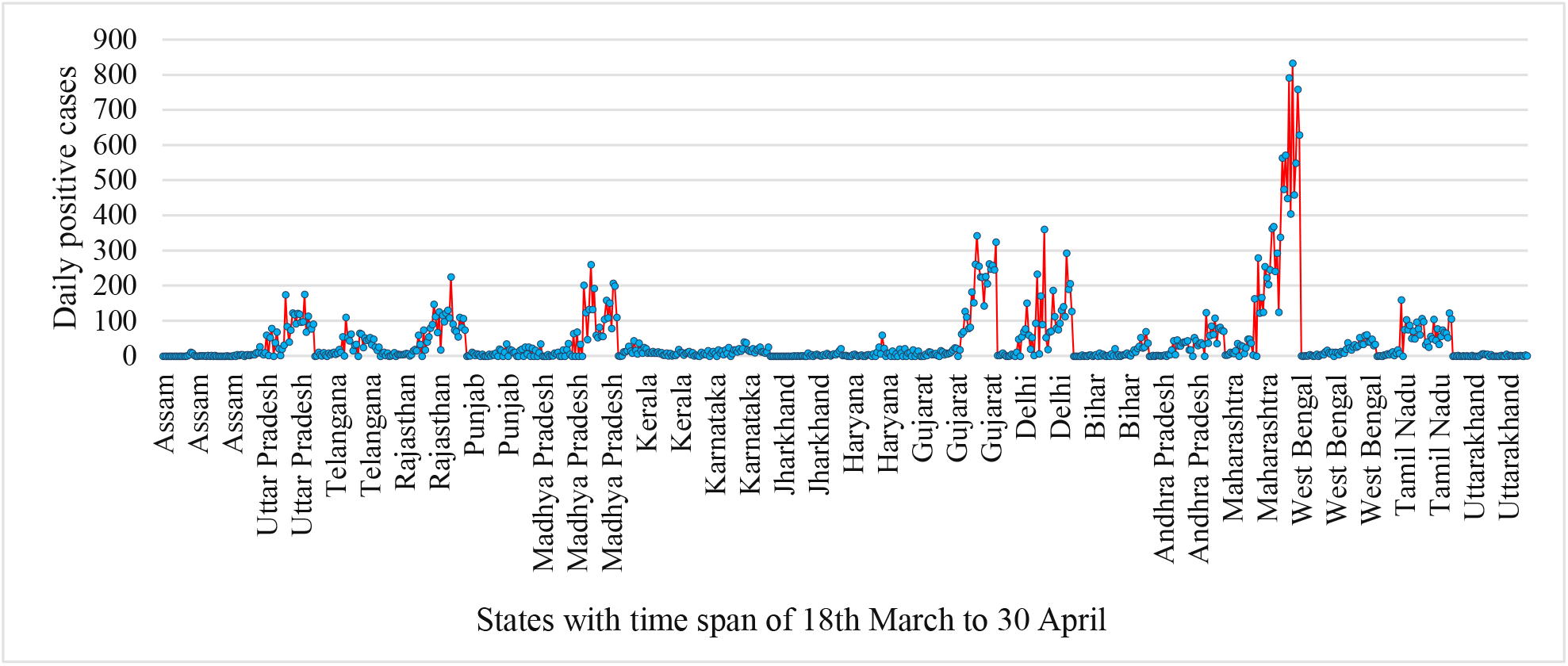
State wise daily COVID-19 positive cases from 18th March to 30th April, 2020.

Table 2 shows Spearman’s correlation coefficients among the COVID case, metrological variables and pollutants. Where temperature is positively correlated with CO (r=0.328p<0.05), PM_2.5_(r=0.108, p<0.05) and COVID-19 cases (r=0.404, p<0.05). However with wind speed (r=0.248, p<0.05) and rainfall (r=-0.148, p<0.05) mean temperature share a low negative relationship. With daily reported corona positive case all variables share a low to moderate negative relationship with 95% confidence level, except temperature whereas wind speed and carbon monoxide (CO) are not significant.

**Table 2.** Spearman’s correlation matrix

| Variables | PM2.5 | PM10 | NO2 | SO2 | CO | WS | SR | RH | Temp | RF | case |
| --- | --- | --- | --- | --- | --- | --- | --- | --- | --- | --- | --- |
| PM2.5 | **1** | **0.659** | **0.521** | **0.418** | **0.348** | **-0.181** | -0.028 | **-0.418** | **0.108** | **-0.099** | **-0.148** |
| PM10 | **0.659** | **1** | **0.452** | **0.546** | **0.360** | -0.041 | -0.036 | **-0.325** | 0.000 | -0.040 | **-0.214** |
| NO2 | **0.521** | **0.452** | **1** | **0.165** | **0.106** | **-0.175** | 0.046 | **-0.191** | -0.055 | **-0.136** | **-0.084** |
| SO2 | **0.418** | **0.546** | **0.165** | **1** | **0.424** | **0.227** | 0.028 | **-0.247** | **-0.072** | **0.152** | **-0.320** |
| CO | **0.348** | **0.360** | **0.106** | **0.424** | **1** | **-0.094** | **-0.313** | **-0.346** | **0.328** | -0.001 | -0.021 |
| WS | **-0.181** | -0.041 | **-0.175** | **0.227** | **-0.094** | **1** | **0.204** | -0.038 | **-0.248** | **0.157** | -0.055 |
| SR | -0.028 | -0.036 | 0.046 | 0.028 | **-0.313** | **0.204** | **1** | -0.022 | **-0.187** | **0.071** | **-0.208** |
| RH | **-0.418** | **-0.325** | **-0.191** | **-0.247** | **-0.346** | -0.038 | -0.022 | **1** | **-0.261** | **0.294** | **-0.070** |
| Temp | **0.108** | 0.000 | -0.055 | **-0.072** | **0.328** | **-0.248** | **-0.187** | **-0.261** | **1** | **-0.148** | **0.404** |
| RF | **-0.099** | -0.040 | **-0.136** | **0.152** | -0.001 | **0.157** | **0.071** | **0.294** | **-0.148** | **1** | **-0.168** |
| case | **-0.148** | **-0.214** | **-0.084** | **-0.320** | -0.021 | -0.055 | **-0.208** | **-0.070** | **0.404** | **-0.168** | **1** |

Values in bold are different from 0 with a significance level $\alpha=0.05$

### 3.2 Effect of weather variables and pollutants on transmitting COVID-19 confirmed case

Firstly in order to identify optimum number of variables associated with the daily case number of COVID-19, a step wise linear regression has been set up with least value of AIC by (lm) function of R software. With the 0-10 days dataset, initial step of model when only intercept was include the value of AIC was 643.04 and at the finishing step it showed lower AIC value 602.26 by including day, temperature, solar radiation and rainfall because of COVID-19 cases are concentrated in some states only. However, 0-25 days dataset indicates AIC with 3091.16 having quite different result includes day, SO_2_, temperature and wind speed. Whereas 0-44 days datasets include more number of variables like day, temperature, relative humidity, PM_2.5_, PM_10_, NO_2_, SO_2_, wind speed, solar radiation and rainfall with lowest AIC 6914.56. Dataset of 44 days includes all variables indicated by 25 days datasets because of substantial number sample has been allowed to have some kind of positive or negative relationship.

#### 3.2.1 Weather parameters and COVID-19 cases

Based on the AIC result, GAM model produced exposer-response curve in Fig. 5a, Fig. 5b and Fig. 5c on seeding dataset of 0-10 days, 0-25 days and 0-44 days, showed temperature is the only meteorological variable positively correlated with COVID-19 positively in short and long run with 14 days moving average. Specifically in 10 days study (Fig. 5a) exposer-response curve (p=<0.000) shows no relation in the range of <23°C and become positively linear after the threshold of 23°C. Fig. 5b shows same result of significant (p=<0.0008) nonlinear relationship with slightly negative in < 23°C and becomes positively linear in >23°C. Fig 5c shows two knots of 23°C and 31°C where <23°C characterizes flat and 23°C - 31°C temperature shows a positive linear relation and after 31°C indicates again flat relation with the number of daily counts of COVID19 cases. After removal of Maharashtra state from dataset (18^th^ March – 30^th^ April), in sensitive analysis, the exposer-response curve is showing a linear pattern of correlation with positive cases (Fig. 6).

**Fig 5.**
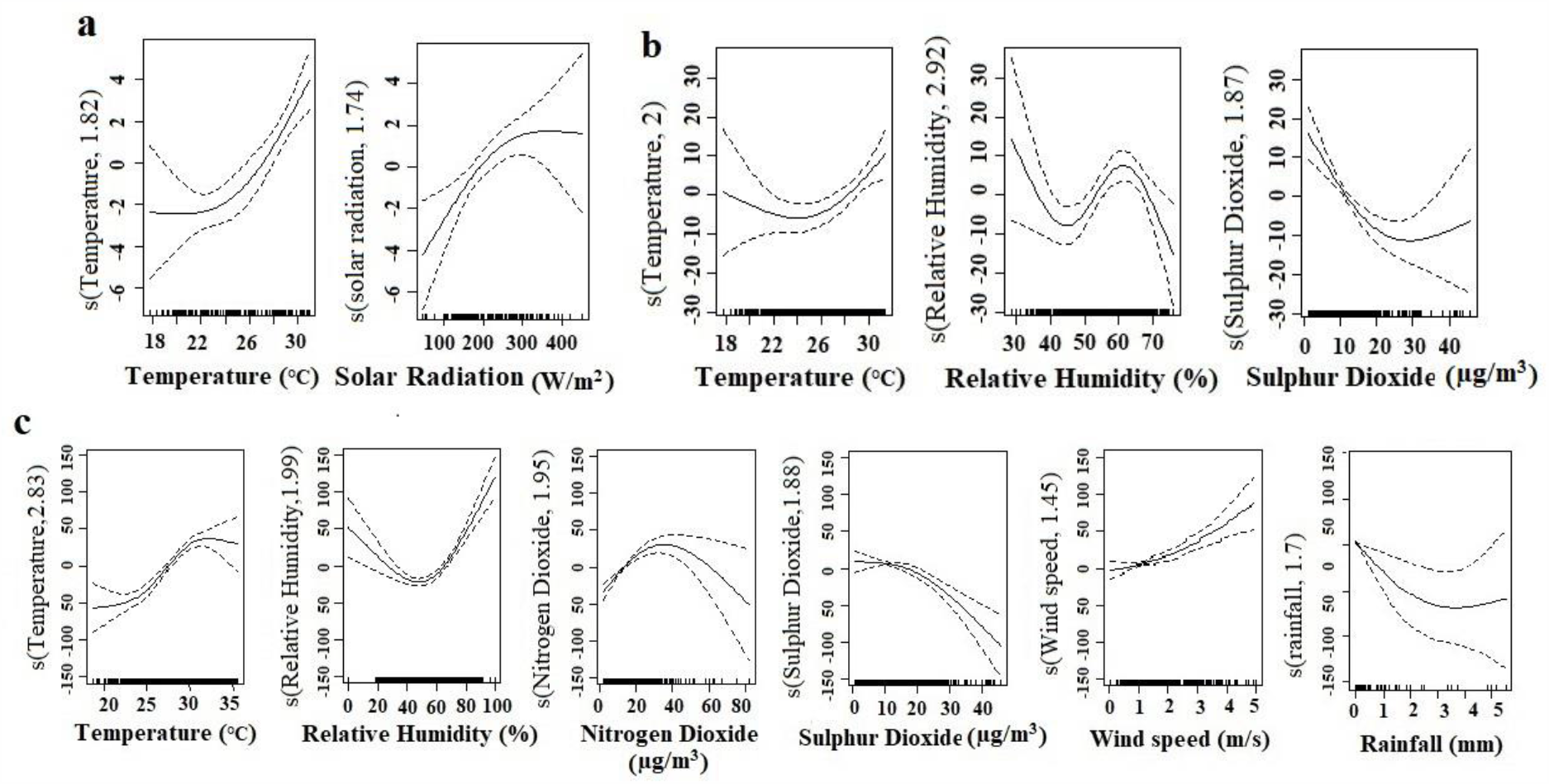
Exposer-Response curve for significant effect of weather and pollutant variables over COVID-19 positive case. The Y axis indicate smoothness threshold to its fitted values and x axis represents mean value of respective variables with 14 days moving average. (a) Result of 0-10 days dataset, (b) result of 0-25 days dataset, (c) result of 0-44 days dataset.

**Fig 6.**
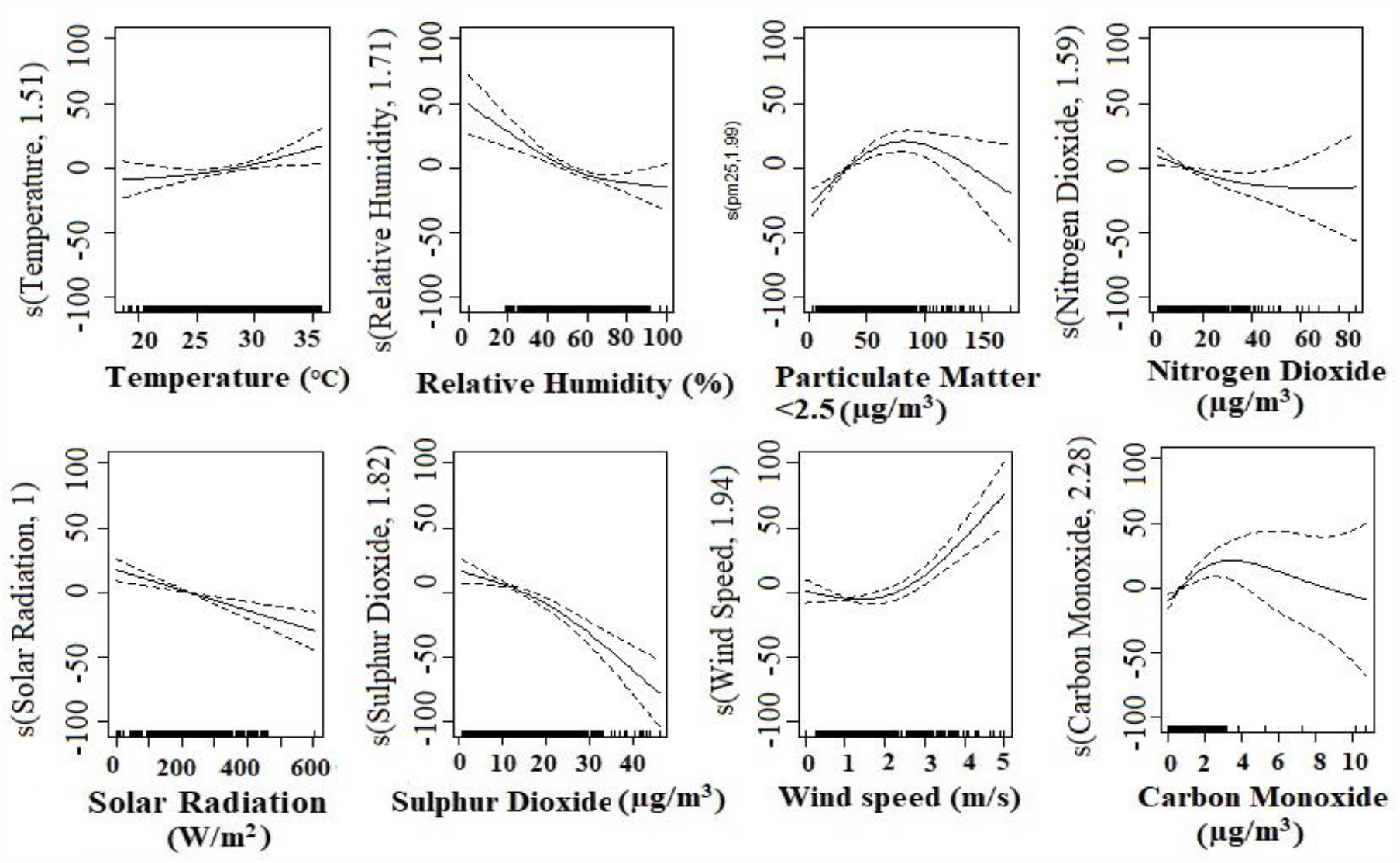
Exposer-Response curve for significant effect of weather and pollutant variables over COVID-19 positive case. The Y axis indicate smoothness threshold to its fitted values and x axis represents mean value of respective variables with 14 days moving average of 0-44 datasets after excluding Maharashtra state.

There is no significant relationship of relative humidity has been found with COVID-19 case in a 10 days span. It may be due to insufficient number of cases were included in some states. But in 25 days of study it was showing negative trend between 30 – 45%, a positive linear trend between 45 – 60% and again negative trend in >60%. 44 days of data set represent a negative trend between 0 – 45% and a positive correlation >45%. But after sensitive analysis it denoted a sharp negative trend all over the curve. The exposer-response curve in Fig. 5c wind speed showed a significant nonlinear positive relationship whereas in spearman’s correlation it was not significant. Wind speed between 1 to 2 m/s shows no relationship curve and >5m/s displayed a positive relation which is contrary of many research work (Ahmadi et al., 2020). In that case when we repeat the model by excluding Maharashtra, it has shown wind speed insignificant. Solar radiation in short period it could have effect on COVID-19 cases but in 44 days of study we have found insignificant relationship. Tough rainfall occurs in very limited areas with maximum 5.53mm with a mean of 0.096mm (Table 1). Even though in 44 days of study it showed a negative relationship. After sensitive analysis rainfall became insignificant, solar radiation became significant and found a negative linear relation. Wind speed maintained a flat trend with the cases in most of the states having mean wind speed in between 0 - 2.5 m/s (Fig. 6).

#### 3.2.2 Pollutants and COVID-19 cases

In fig. 5 we have not found any significant relation of PM_2.5_, PM_10_, NO_2_, SO_2_ and CO with COVID-19 cases in 10 days study. But in 25 days there was only SO_2_ with decrease trend with slight positive after concentration of 35 µg/m^3^. A significant negative relationship of SO_2_ with COVID-19 cases has been recorded. It showed 0-10 mg/m^2^ have flat and followed by a step decreased trend in concentration between 10-45 µg/m^3^. NO_2_ has a significant positive correlation in between 0-35 µg/m^3^ and followed by a decrease trend after > 40 µg/m^3^ (Fig. 5c).Again it displayed a fresh negative correlation during sensitive analysis. In case of PM_2.5_ and PM_10_ it shows significant correlation with 6 degree of freedom therefore because of overfitting we eliminate it (Zhu et al., 2020 and Wang et al., 2018). After sensitive analysis there is huge change appeared in association of pollutants in GAM model. PM_2.5_, and CO have become significant (Fig 6). There was a positive relationship with PM_2.5_ showed through exposer-response curve, in most of the states, up to the concentration of 100 µg/m^3^. Same pattern followed by carbon monoxide (CO).

### 3.3. Sensitive Analysis

GAM result on 44 days data including Maharashtra state displayed Temperature, Relative humidity, NO_2_, SO_2_, wind speed and rainfall (Estimate 42.07, SD 2.66 and *p*<2e-16) as significant. For sensitive analysis, first we excluded Maharashtra state having a large number of case concentrate in last couple of days (Fig. 4) than other state for robust result (Zhu et al., 2020). Fig. 6 present a robust study output (Estimate 30.75, SD 1.468 and *p*<2e-16) where PM_2.5_, solar radiation and CO along with temperature, relative humidity, NO_2_ SO_2_ and wind speed have shown a significant association with COVID-19. With the help of this scenario it can be stated that pollutants like NO_2_, PM_2.5_, PM_10_ and CO are more sensitive variables than SO_2_ in this study. Weather variables like relative humidity and solar radiation and rainfall are more sensitive than temperature and wind speed.

## 4. Discussion

Pandemic COVID-19 became burden and its prolonged impact in every sphere of human life could be sensed even after successive decades. Our effort is to find out the association meteorological factors and pollutants with daily positive case of COVID-19. GAM model with 14 days moving average of temperature and showed a significant positive relationship with temperature, relative humidity, and wind speed whereas Relative Humidity, NO_2_, SO_2_, and rainfall and solar radiation showed a negative relation with daily case in 44 days of study. With the help of previous research work (Kumar, 2020 and Ahmadi et al., 2020) shows a positive and negative relation with temperature and relative humidity respectively. Similar to Chan et al., 2010, our study also shows high temperature leads to minimise the reported case. But it will need more experiment to establish a strong relationship with temperature. In the case of relative humidity two type of scenario has been seen. Firstly, landlocked States like Rajasthan, Madhya Pradesh, Uttar Pradesh Bihar, Punjab, and Himachal Pradesh (RH <57%) have shown a negative association with daily positive case. Secondly, Coastal states like Maharashtra, Kerala, Tamil Nadu and Andhra Pradesh with high relative humidity (>57%) having a positive association with cases. After excluding Maharashtra It seems a negative correlation in between 0 - 90%, which is again validated by previous study (Kumar, 2020, Qi et al., 2020 and Ma et al., 2020). Again relative humidity may be statistically significant till it need more research to establish as a fact. In a study in Asyary et al., 2020 claim that Sunlight correlated significantly to recovery patient of COVID-19 because Vitamin D from sunlight can triggered body immune system. In our study we have seen a negative relation between infected cases with sunlight exposer. There are states like Maharashtra, Kerala, Rajasthan Haryana, Delhi and Punjab enjoy solar radiation more as reported cases are also more. Therefore solar radiation will not be a establish factors can reduce COVID-19 transmission (Gupta et al., 2020). Wind speed again pointed out a positive correlation can be validate with previous study in India (Gupta et al., 2020). Many study found (Xie et al., 2019, Tecer et al., and 2008 and Lu et al., 2015) respiratory disease like asthma and respiratory infection have a significant association with atmospheric particulate matter. Chen et al., 2007 established a positive relationship with exposer of SO_2_, NO_2_ and CO with respiratory disease. In our study, we found PM_2.5_ and CO positively associated with transmission of positive cases where as NO_2_ and SO_2_ have a negative relation it is because of lockdown where most of the factories and transportation were suspend. This study also predict Gujrat, Uttar Pradesh, Rajasthan, Madhya Pradesh, West Bengal, Delhi and Bihar have more concentration of particulate matter that may enhance more serious respiratory disease as well as COVID-19 epidemic.

This study has many limitations like not inclusion of socio-cultural factors, gender, population density, movement of people and elevation etc. There is several factors can help to transmit this disease. Further laboratory and analytical studies are needed on these shortcomings.

## 5. Conclusion

Our findings are there is a significant relationship between daily positive COVID-19 case with weather and pollution factors. Temperature and wind speed is positively associated with daily COVID-19 case count. Solar radiation has negative correlation. Positive association has been found PM_2.5_ and CO with COVID-19 confirmed cases and negative association with SO_2_ and NO_2_. Besides environmental factors, socio-economic factors, health infrustcture, literacy, number of person came from abroad, number of suspects in a states and their travel history can also enhance the accuracy of understanding actual influential factors in future study.

We have no conflict of interest of this paper.

## Data Availability

For data, one can contact me in

https://orcid.org/0000-0001-9948-1577

